# Bias-adjusted predictions of county-level vaccination coverage from the COVID-19 Trends and Impact Survey

**DOI:** 10.1101/2022.05.18.22275217

**Authors:** Marissa B. Reitsma, Sherri Rose, Alex Reinhart, Jeremy D. Goldhaber-Fiebert, Joshua A. Salomon

**Author notes:** **Correspondence to:** Marissa Reitsma, 615 Crothers Way Encina Commons, Stanford University, Stanford, CA 94305.

## Abstract

The potential for bias in non-representative, large-scale, low-cost survey data can limit their utility for population health measurement and public health decision-making. We developed a multi-step regression framework to bias-adjust vaccination coverage predictions from the large-scale US COVID-19 Trends and Impact Survey that included post-stratification to the American Community Survey and secondary normalization to an unbiased reference indicator. As a case study, we applied this framework to generate county-level predictions of long-run vaccination coverage among children ages 5 to 11 years. Our vaccination coverage predictions suggest a low ceiling on long-term national coverage (46%), detect substantial geographic heterogeneity (ranging from 11% to 91% across counties in the US), and highlight widespread disparities in the pace of scale-up in the three months following Emergency Use Authorization of COVID-19 vaccination for 5 to 11 year-olds. Generally, our analysis demonstrates an approach to leverage differing strengths of multiple sources of information to produce estimates on the time-scale and geographic-scale necessary for proactive decision-making. The utility of large-scale, low-cost survey data for improving population health measurement is amplified when these data are combined with other representative sources of data.

## Background

The COVID-19 pandemic highlighted the importance of local and timely indicators to inform public health decision-making, but such indicators have remained elusive in areas critical to pandemic response. For example, indicators of people’s vaccination intentions could ideally be used to predict subsequent vaccine uptake and to drive targeted efforts to reduce hesitancy and thereby increase achieved coverage. However, representative survey data are too costly to collect repeatedly with samples large enough for county-level estimation in the United States, while unrepresentative large-scale survey data have been shown to yield biased estimates with misleadingly small margins of error.^1–3^ Although programmatic data offer retrospective reporting of coverage at county-level, these data become available too late to enable prospective planning and decision-making, and many important indicators do not have routine reporting systems.^1,4^

Combining data sources with different advantages and limitations can help to balance tradeoffs between time, cost, and representativeness of data collection. Studies in other areas of health have combined multiple data sources for retrospective bias correction and small area estimation.^5–8^ The COVID-19 pandemic catalyzed a new era of massive real-time data collection for public health, exemplified by the US COVID-19 Trends and Impact Survey, which has been running daily since April 2020.^2^ The US survey has an average of 40,000 responses daily. Its size allows for timely small-area estimation of many policy-relevant leading indicators, but its utility has been questioned due to bias in estimates of vaccination coverage compared to representative reporting data.^3^ Approaches to gain actionable insights from these large-scale survey data are relevant to current COVID-19 pandemic response, and to general population health measurement, for which similar large-scale, low-cost surveys could be deployed in the future.

Although COVID-19 vaccination has been central to the public health response to the pandemic, coverage has plateaued well below 100%, with wide variation across communities. COVID-19 vaccination intentions have been an important indicator derived from survey data over the course of the pandemic.^5,9–13^ Vaccination intentions can be used to anticipate eventual vaccination coverage for different groups, which can then be used to direct resources and targeted interventions, design policies, deploy additional risk reduction tools, and monitor both the pace and equity of scale-up. In the United States, children ages 5 to 11 years became the most recently eligible group for COVID-19 vaccination when Emergency Use Authorization was extended at the end of October 2021.^14^ In this study, we present a framework to bias-adjust estimates of vaccine intentions from the large-scale COVID-19 Trends and Impact Survey and predict future county-level vaccination coverage plateaus, using vaccination among children ages 5 to 11 years as an illustrative case study.

## Methods

We developed a multi-step regression framework (Figure 1) to predict vaccination coverage plateaus among children ages 5 to 11 years. First, we estimated county-level parental hesitancy toward vaccinating their children using a mixed effects logistic regression model fit to survey data. Next, we estimated the relationship between county-level hesitancy and observed vaccination coverage, for youth ages 12 to 17 years, who became eligible for COVID-19 vaccination earlier than children ages 5 to 11 and therefore provide a reference group, using a second logistic regression model. Finally, we combined the results from the two regression models to predict county-level vaccination coverage for 5- to 11-year-olds.

**Figure 1.**
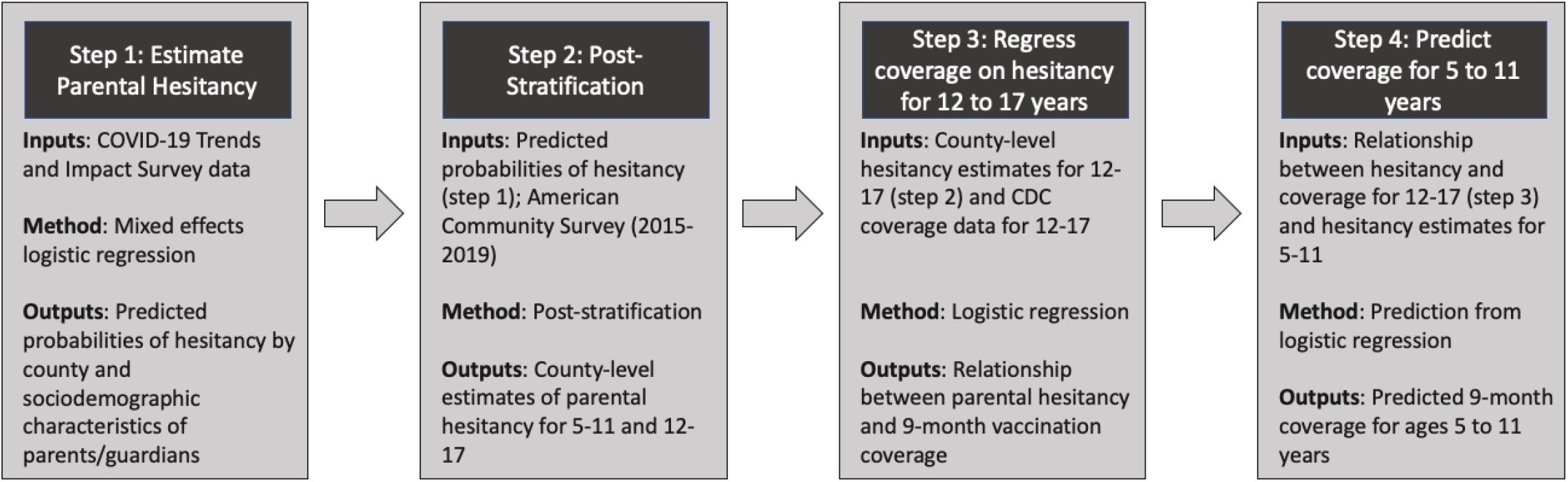
Methods Flowchart.

### Data Sources

We combined individual-level survey responses from Wave 11 of the COVID-19 Trends and Impact Survey (CTIS) collected during the period from July 1, 2021, through October 31, 2021, with data from Wave 12, collected during the period from December 19, 2021, through February 14, 2022. The survey is managed and implemented by the Delphi Group at Carnegie Mellon University. Participants are recruited through Facebook and the sampling frame is the Facebook Active User base. Additional information on the COVID-19 Trends and Impact Survey has been previously published.^2^ The full questionnaire for Waves 11 and 12 is available online.^15^

In addition to CTIS, we used individual-level sociodemographic data (age, documented sex, education, race/ethnicity, and household structure) from the 2015-2019 American Community Survey for post-stratification.^16^ Individual-level data from the American Community Survey are available at the public-use microdata area level. We mapped public-use microdata areas to counties using the Missouri Census Data Center’s Geographic Correspondence Engine.^17^ When a single county contained multiple public-use microdata areas, we aggregated public-use microdata areas to the county-level. When a single public-use microdata area spanned multiple counties, we assumed the same distribution of sociodemographic characteristics for each county.

Finally, we used complete vaccination coverage data for ages 12 to 17 years reported at the county-level by the Centers for Disease Control and Prevention for the second-stage regression, and data from the same source over the first three months after eligibility for ages 5 to 11 years for performance evaluation of coverage predictions.^18^

### Estimating County-Level Hesitancy

To estimate county-level parental hesitancy, we fit a mixed-effects logistic regression to survey data on attitudes of parents/guardians towards vaccinating their children. We classified “No, definitely not” and “No, probably not” as hesitant responses to the question “Will you choose to get a COVID-19 vaccine for your child or children when they are eligible?”. Responses of “Yes, definitely” and “Yes, probably” were considered not hesitant. Consistent with previous analyses, we used the imprecise but available construct of “reported hesitancy” and focused on it principally as an intermediate indicator that would be subsequently mapped to long-run vaccination coverage.

The CTIS survey questionnaire evolved as new information became available over the course of the pandemic. Importantly, while Wave 11 asked parents about vaccine hesitancy, it did not ask for the ages of their children. Since Wave 12 elicited the age of the parent’s oldest child, we used it to examine differences in parental hesitancy for those whose oldest child was between the ages of 12 and 17 versus ages 5 to 11.

The first-stage logistic regression modeled the probability of parental hesitancy as a function of fixed effects for documented gender (male, female), age group (18-24, 25-34, 35-44, 45-54, 55-64, 65+), education (high school or fewer years of education, some college or a two-year degree, four-year degree, graduate degree), and race/ethnicity (Hispanic, non-Hispanic American Indian or Alaska Native, non-Hispanic Asian, non-Hispanic Black, non-Hispanic Native Hawaiian or Other Pacific Islander, non-Hispanic White, non-Hispanic multiracial or other race), and age group of child (unknown, 12 to 17, and 5 to 11), and nested random intercepts on state and county:

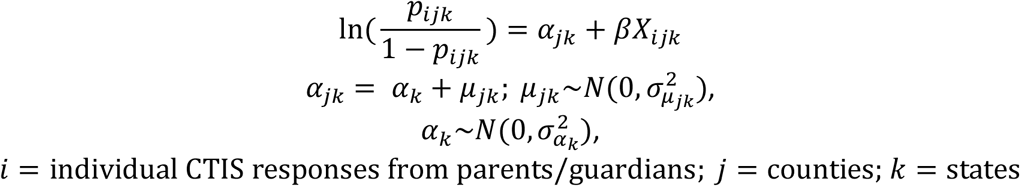

We did not perform a weighted regression to include the CTIS survey weights, instead adjusting for the probability of inclusion and non-response through post-stratification.^19^ We combined data from counties with a sample size of 10 or fewer into grouped counties, by state. To generate county-level predictions of hesitancy, including uncertainty around these predictions, from the first-stage regression, we generated 1,000 draws of subgroup-level predicted probabilities of hesitancy for unique combinations of documented gender, age group, education, race/ethnicity, and county using the estimated regression coefficients, assuming a multivariate normal distribution of the parameters including the fixed and random effects 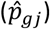, where *g* corresponds to each unique demographic characteristic combination. We then post-stratified county- and subgroup-level predicted probabilities of hesitancy to produce overall county-level hesitancy estimates 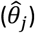:

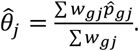

The weights (*w*_*gj*_) used for post-stratification were based on an analysis of individual-level data from the American Community Survey that reflected household structure and incorporated children’s sample weights. First, we identified each child’s parents/guardians based on the first available of the following: 1) parents directly coded in the American Community Survey, 2) grandparents designated as responsible for one or more children directly coded in the American Community Survey, 3) adults (18+) in the same household and same family unit, and 4) adults (18+) in the same household but different family unit. Next, we assigned the child’s sample weight to each of their parents/guardians, dividing the weight by the total number of identified parents/guardians. Finally, we summed the children’s sample weights across each subgroup (*g*), defined by the demographic characteristics of the parents/guardians, and each county (*j*), resulting in the final weight (*w*_*gj*_) used for post-stratification.

### Predicting County-Level Vaccination Coverage from Hesitancy Estimates

We used a second logistic regression model to translate county-level hesitancy estimates to county-level vaccination coverage predictions. We trained the model on paired county-level hesitancy and coverage estimates for children ages 12 to 17 years, and then projected the predictive relationships onto the hesitancy estimates for children ages 5 to 11 years under the assumption that the same relationships would apply across the two age groups. To estimate the regression model for the 12 to 17 year group, we first generated estimates of parental hesitancy for this group using the same regression model specification described above for the 5 to 11 year group, in this case predicting hesitancy for parents of children ages 12 to 17 years and post-stratifying estimates based on household structure and sample weights of children ages 12 to 17 years. These county-level hesitancy estimates were used as independent variables in the second logistic regression. For our dependent variable, we used coverage data from February 1, 2022, which was approximately nine months after 12-17 year-olds first became eligible for vaccination (ages 16-17 years in early April 2021 and ages 12-15 years on May 10, 2021).

For states with at least ten counties reporting vaccination coverage data for children ages 12 to 17 years on February 1, 2022, with CDC reporting completeness exceeding 80%, we fit state-specific regressions. For all other states and the District of Columbia (n=7), we fit regressions at the census division level. This prevented overfitting to small numbers of counties or low-quality reporting data. Regressions were weighted based on the size of the 12 to 17 population in each county. The second regression was fit to each of the 1,000 draws of county-level hesitancy from the first regression. Uncertainty from the second regression was captured through 1,000 draws from the multivariate normal distribution of the fixed effects plus the residual variance.

Finally, we used the models fit on the relationship between parental hesitancy and vaccination coverage for children ages 12 to 17 to predict coverage for children ages 5 to 11 years based on our first-stage estimates of hesitancy for this age group. Final prediction intervals were based on the 2.5^th^ and 97.5^th^ percentiles of one million final county-level coverage predictions (1,000 draws from the first regression crossed with 1,000 draws from the second regression).

### Performance Evaluation

We compared our estimates of parental hesitancy towards vaccinating children ages 12 to 17 years to estimates on the same indicator produced by the Office of the Assistant Secretary for Planning and Evaluation (ASPE), including comparing correlation coefficients between estimated county-level hesitancy and observed vaccination coverage on February 1, 2022.

To evaluate our use of the relationship between hesitancy and coverage for children ages 12 to 17, applied to children ages 5 to 11, we assessed interim coverage predictions for the 5 to 11 age group at three months after EUA expansion against observed county-level coverage reported by the CDC, based on the intraclass correlation coefficient and percentage of counties for which the 95% prediction interval contained the observed coverage level nationally and at the state level. Since the CDC does not report separate county-level coverage estimates for ages 12 to 15 versus 16 to 17, the time since an age group first became eligible for vaccination is an imprecise but best-available approach to this interim performance evaluation.

### Monitoring Progress and Equity in Scale-Up

To monitor pace of vaccination scale-up, we defined a measure of “3-month progress” as:

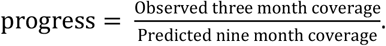

To monitor equity in the pace of vaccination scale-up, we used linear regression to analyze associations between this progress measure and the county-level socioeconomic status domain of the CDC’s Social Vulnerability Index, which reflects measures of poverty, unemployment, income, and education.^20^

### Study Approval and Data Availability

The study was approved by Stanford’s Institutional Review Board, under protocol number 56018. All analyses were conducted using the R programming language version 3.6.3. Analytic code is available through GitHub (https://github.com/PPML/CTIS-County-Vaccination-Coverage). The COVID-19 Trends and Impact Survey microdata can be accessed through a data use agreement with Carnegie Mellon University, while the American Community Survey data and CDC vaccination data are publicly available.

## Results

### Data

Between July 1 and October 31, 2021, a total of 613,460 responses to Wave 11 of the US COVID-19 Trends and Impact Survey (CTIS) were collected from parents/guardians of children under age 18 with complete demographic information. To allow for variation in parental hesitancy by child age group, we supplemented the analyses with 119,465 responses collected from parents/guardians whose oldest children were between the ages of 5 and 17 years in Wave 12, between December 19, 2021 and February 14, 2022. We report exclusions in the sample flowchart (Supplemental Figure S1). Unweighted and weighted distributions of respondents by age, documented gender, education, and race/ethnicity are reported in Table 1. Post-stratification to the American Community Survey reduced bias from the non-representative sample. Of 3,142 counties, 1,203 had a sample size of at least 100, while 293 had a sample size between one and 10, and 23 had zero respondents. Maps of county-level sample sizes and sample rates are reported in Supplemental Figures S2 and S3.

**Table 1.** Unweighted and weighted distribution of sociodemographic variables of included guardians from the COVID-19 Trends and Impact Survey (CTIS), compared to distribution of sociodemographic variables of parents/guardians in the American Community Survey (ACS)

|  | CTIS Survey<br>Data:<br>Unweighted | CTIS Survey<br>Data:<br>Weighted | Parents/Guardians<br>of Children Ages<br>12-17 | Parents/Guardians<br>of Children Ages<br>5-11 |
| --- | --- | --- | --- | --- |
| **Documented Gender\*** |  |  |  |  |
| Female | 65.4% | 53.9% | 55.4% | 55.4% |
| Male | 34.6% | 46.1% | 44.6% | 44.6% |
| **Age Group** |  |  |  |  |
| 18-24 | 1.9% | 5.8% | 0.7% | 1.4% |
| 25-34 | 13.7% | 17.7% | 8.4% | 29.6% |
| 35-44 | 29.0% | 28.3% | 42.1% | 48.7% |
| 45-54 | 25.6% | 24.4% | 39.3% | 16.8% |
| 55-64 | 16.0% | 13.6% | 7.9% | 2.5% |
| 65+ | 13.7% | 10.2% | 1.6% | 1.0% |
| **Education** |  |  |  |  |
| High school or less | 21.7% | 25.1% | 37.1% | 35.9% |
| Some college or two-year degree | 35.9% | 35.9% | 29.7% | 29.8% |
| Four-year degree | 23.2% | 21.7% | 20.4% | 20.7% |
| Graduate degree | 19.2% | 17.3% | 12.8% | 13.6% |
| **Race/Ethnicity** |  |  |  |  |
| American Indian or Alaska Native | 1.1% | 1.0% | 0.7% | 0.7% |
| Asian | 2.8% | 3.4% | 5.4% | 5.9% |
| Black | 7.1% | 7.2% | 11.5% | 11.5% |
| Hispanic | 17.3% | 21.7% | 22.0% | 23.0% |
| Native Hawaiian or Other Pacific Islander | 0.3% | 0.3% | 0.2% | 0.2% |
| Other | 4.8% | 5.2% | 1.8% | 2.0% |
| White | 66.6% | 61.2% | 58.4% | 56.7% |

\*CTIS collects information on the respondent's self-reported gender, while the ACS collects information on the respondent's self-reported sex.

### Hesitancy Estimates

Modeled county-level parental hesitancy toward vaccination for children ages 5 to 11 years ranged from 7% (95% Prediction Interval: 5-9%) in San Mateo County, California to 74% (61-84%) in Platte County, Wyoming. Although the population-weighted national average hesitancy was 31%, 2,787 counties (89% of all counties) had hesitancy levels exceeding this benchmark. The skewed distribution of county-level estimates versus state- and national-aggregates is largely driven by lower hesitancy in urban areas with large populations and higher hesitancy in rural areas with smaller populations. Across counties, median hesitancy towards vaccination was 21% (IQR: 18-25%) higher for parents of children ages 5 to 11, compared to parents of children ages 12 to 17. Our estimates of hesitancy among parents of children ages 12 to 17 using data from CTIS reflected substantially more sub-state variation in hesitancy, compared to previously published estimates from ASPE (Figure 2). Additionally, our estimates showed a stronger correlation with vaccination coverage on February 1, 2022, among children ages 12 to 17 years (CTIS: -0.78; ASPE: -0.44) (Supplemental Figure S4).

**Figure 2.**
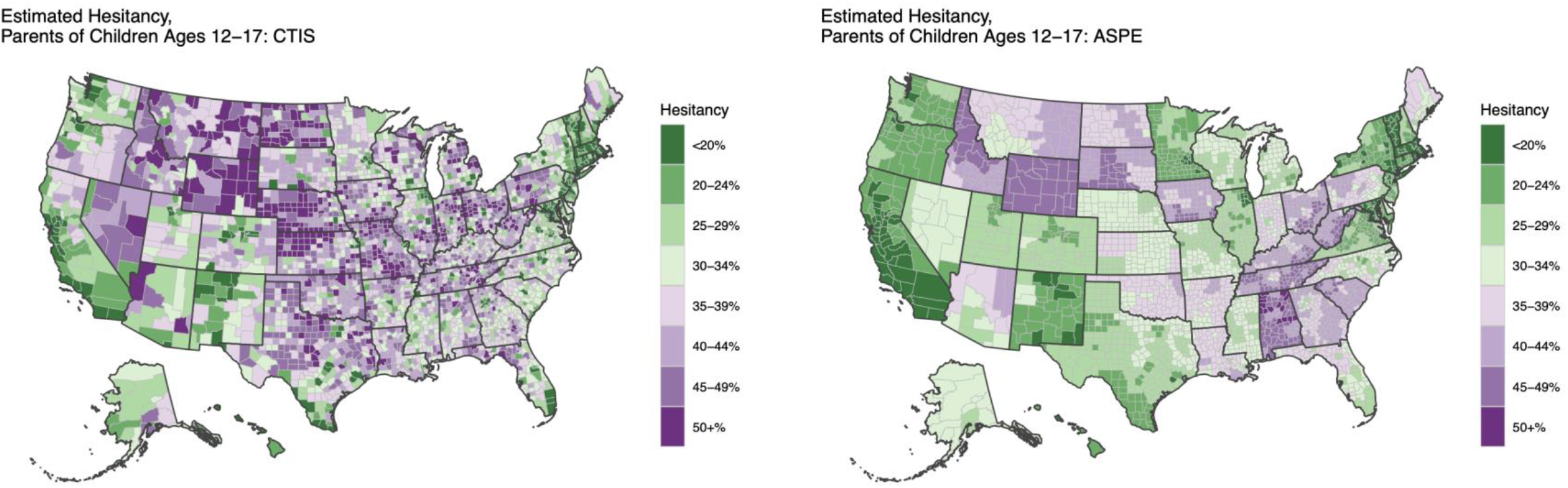
Comparison of county-level hesitancy estimates for parents of children ages 12-17 produced by CTIS (left) and ASPE (right).

### Predicted Coverage Levels

Predicted mean national plateau coverage level among children ages 5 to 11 by August 2022, nine months after EUA, was 46%. There was substantial state-level variation in predicted plateau coverage, ranging from 30% and below in Wyoming, Alabama, Mississippi, Idaho, and Louisiana to 66% and above in Connecticut, District of Columbia, and Massachusetts. Four out of the five counties with the highest predicted coverage were in California. Ninety-two percent of counties are predicted to fall short of a 50% coverage benchmark by August 2022 for children ages 5 to 11, while 56% of counties are predicted to not reach 30% coverage. Eighty-six percent of counties are predicted to fall short of their state average coverage level, highlighting an urban-rural divide in vaccination. Higher levels of predicted coverage are concentrated in the northeast, west coast, and in urban centers across the country (Figure 3).

**Figure 3.**
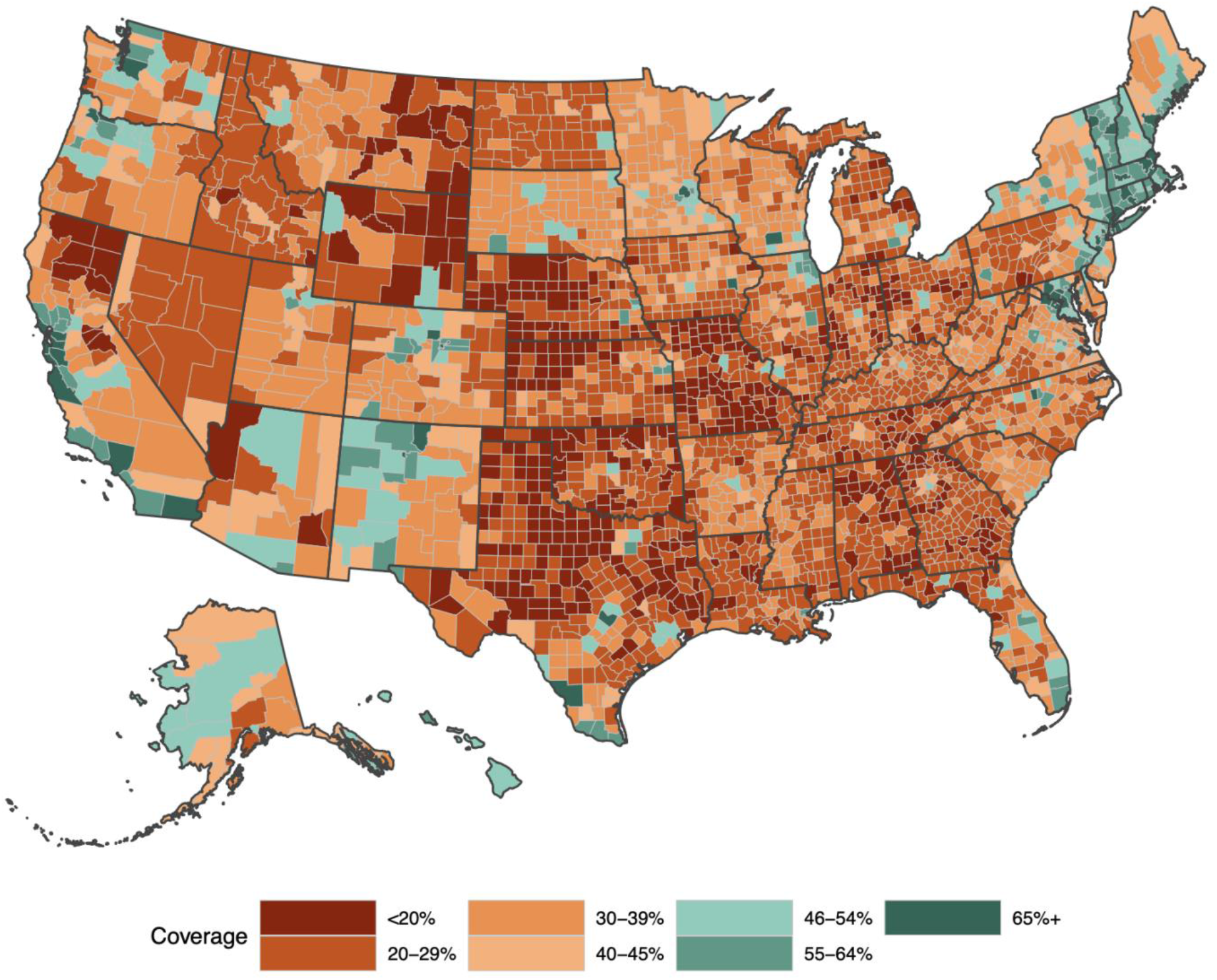
County-level map of predicted plateau complete vaccination coverage levels by August 2022 for children ages 5-11 years. The color scale is split at the national average predicted coverage of 46%.

### Model Validation

Figure 4 shows the relationship between predicted three-month coverage among children ages 5 to 11 years and observed coverage three months after EUA. The intraclass correlation coefficient for consistency of predicted versus observed three-month coverage at national level was 0.81. Intraclass correlation coefficients were greater than 0.75 for 16 states, between 0.50 and 0.75 for 19 states, and less than 0.50 for 10 states. Intraclass correlation coefficients at state level are reported in Supplemental Table S5. The prediction interval for three-month coverage included the observed coverage level for 81% of counties.

**Figure 4.**
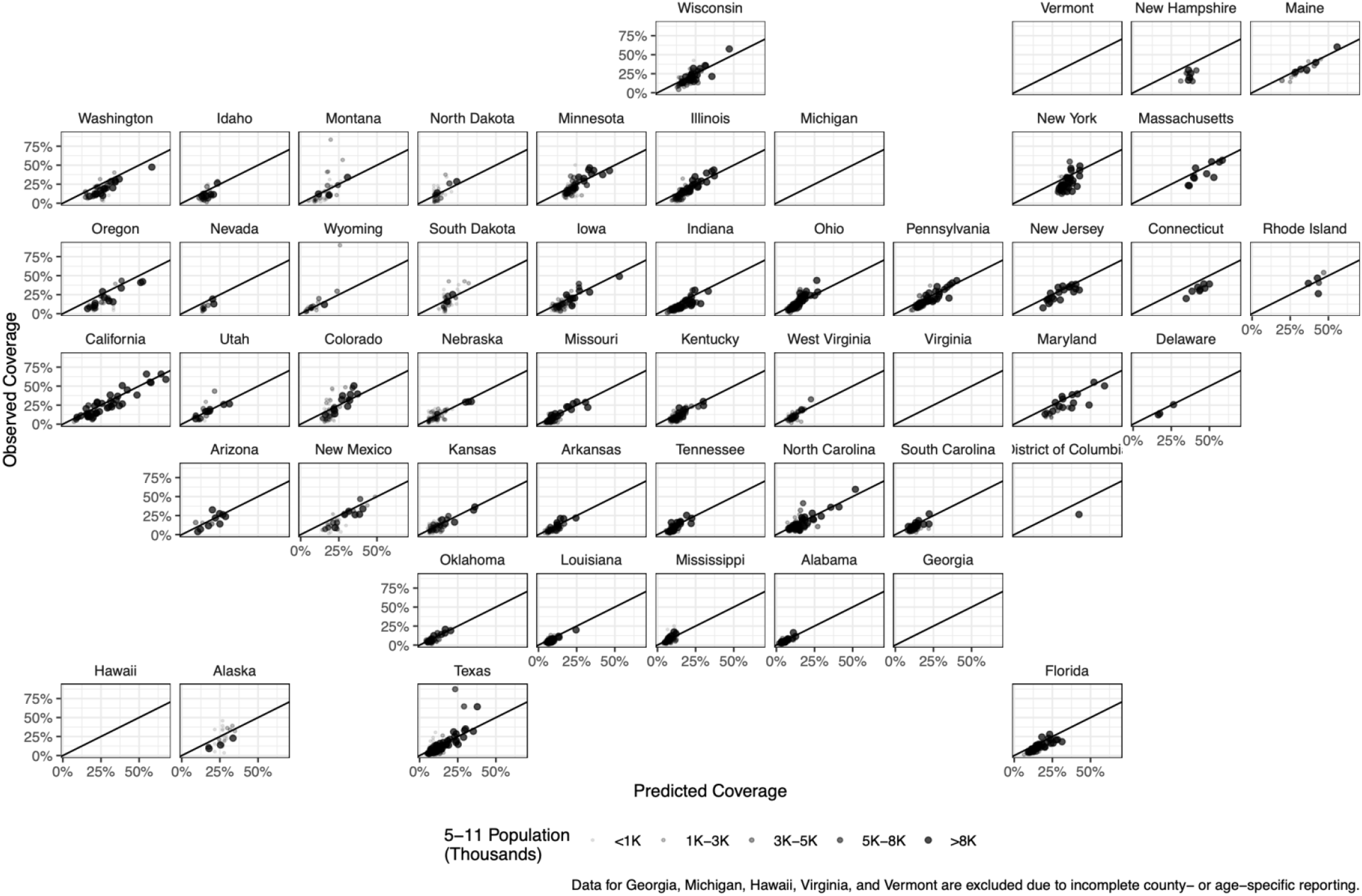
Three-month validation of county-level predicted versus observed complete coverage among children ages 5 to 11 years.

### Monitoring Progress and Equity in Scale-Up

Relative to long-term predicted coverage levels, at the state level, Vermont, Rhode Island, and Maine had the fastest pace of scale-up of coverage among children ages 5 to 11 years at three months after EUA, while Louisiana, Alabama, and Mississippi had the slowest pace of scale-up (Figure 5). We find that errors in nine-month coverage predictions among ages 12 to 17 years were not significantly associated with the socioeconomic status domain of the CDC’s Social Vulnerability Index (SVI) in all states except South Dakota, Nevada, and Montana. As a result, in addition to predicting plateau coverage levels, we can use county-level predicted coverage levels to monitor equity in the pace of vaccination scale-up. More vulnerable counties, as measured by the socioeconomic status domain of the CDC’s SVI generally made less progress toward reaching their plateau coverage levels over the first three months after EUA expansion, compared to less vulnerable counties. There was significantly slower scale-up progress in more vulnerable (higher SVI) counties in 36 out of 46 states reporting data on vaccination coverage among children ages 5 to 11 years (Figure 6).

**Figure 5.**
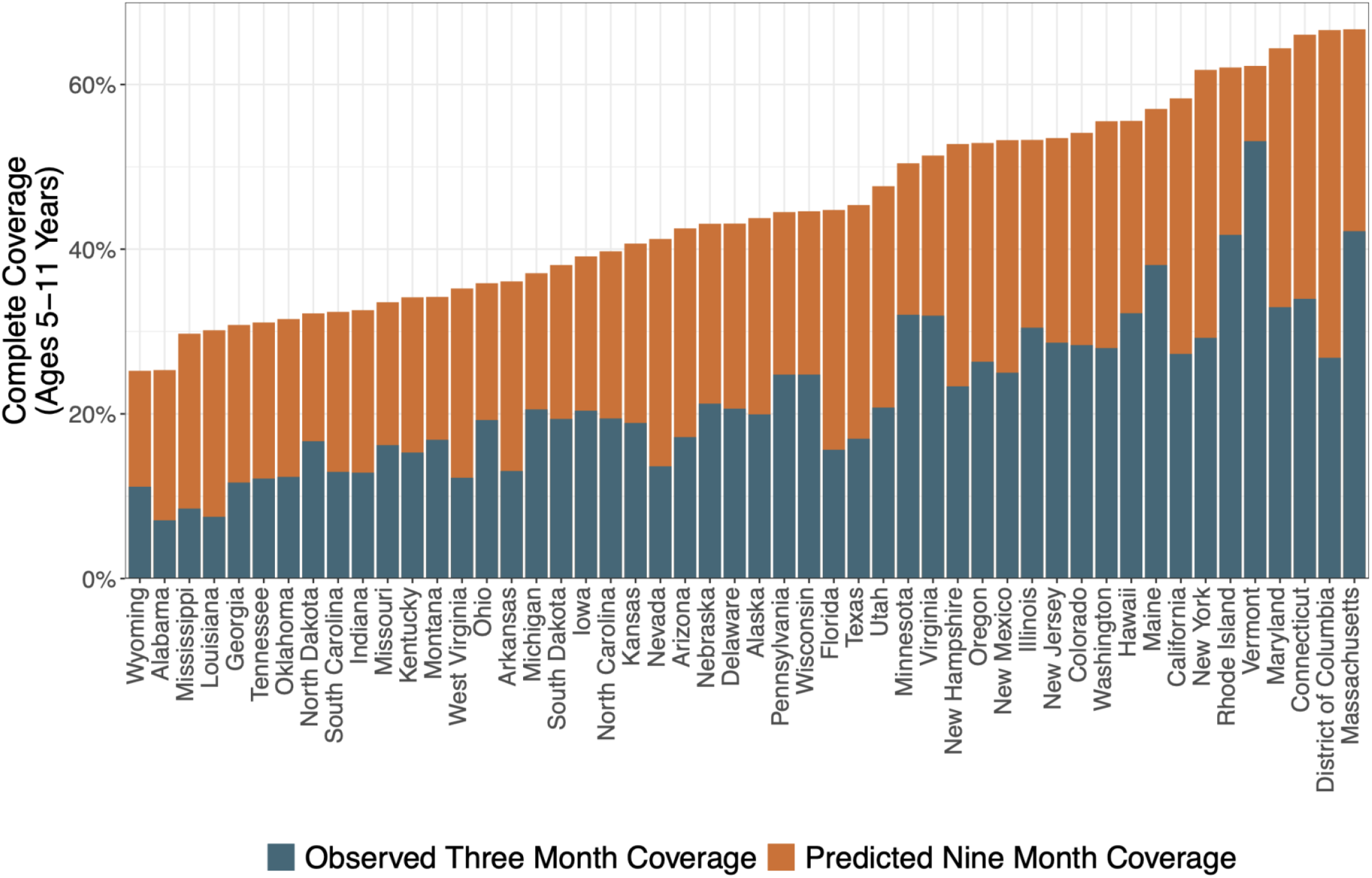
State-level three-month complete vaccination scale-up progress for children ages 5-11 years, and nine-month predicted coverage.

**Figure 6.**
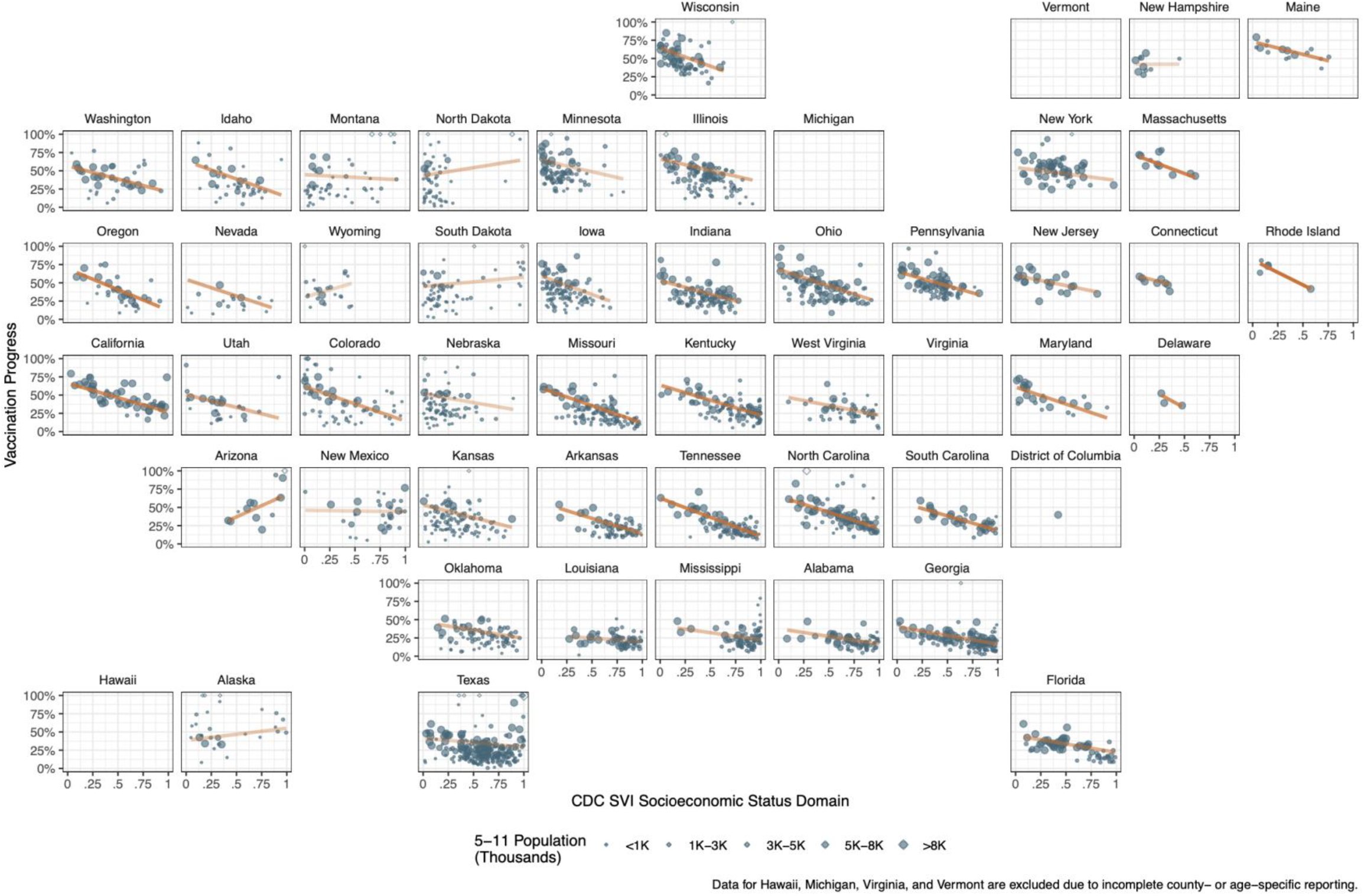
Association between three-month county-level complete vaccination progress for children ages 5-11 years and the socioeconomic status domain of the CDC’s Social Vulnerability Index. The intensity of the trend lines is proportional to the linear regression R^2^.

## Discussion

We generated bias-adjusted county-level predictions of long-term vaccination coverage for children ages 5 to 11 years that leveraged data on parental vaccination intentions from the COVID-19 Trends and Impact Survey (CTIS). To mitigate the impacts of selection bias in the sample, we combined CTIS data with representative sociodemographic data from the American Community Survey and unbiased programmatic data on vaccination coverage. Our approach to estimation and propagation of multiple sources of uncertainty produced prediction intervals that included observed coverage levels for 81% of counties three-months after EUA. Our estimation framework can be broadly used to generate actionable indicators on the time-scale and at the geographic-scale required for decision-making during the pandemic and beyond.

Our predictions for vaccination coverage nine months after EUA for children ages 5 to 11 years suggest that 92% counties are likely to fall short of a 50% coverage benchmark. Across and Within states, there is substantial geographic heterogeneity in both parental hesitancy and predicted coverage. These estimates have implications for targeting of efforts to promote vaccination uptake among eligible children and expectations for eventual vaccination uptake among younger children who are not yet eligible. They also underscore the continued need for other protective measures, including masking, testing, and improved ventilation, in schools during periods of significant community transmission.^21,22^

Despite consistent messaging about the importance of promoting equity in vaccination scale-up, we observe a pervasive pattern of slower vaccination scale-up in more vulnerable counties, as measured by the socioeconomic domain of the CDC’s Social Vulnerability Index.^23–25^ The socioeconomic status domain of the Social Vulnerability Index comprises measures of income, poverty, employment, and education. Moving forward, as vaccination is extended to even younger children and as new rounds of boosters or new vaccines are authorized, more intensive and explicitly pro-equity policies and programs are required to break the cycle of inequity in vaccination scale-up that has been repeated in every phase of the vaccination campaign.^26–29^

Large-scale, low-cost surveys offer a promising approach to population health measurement. They offer advantages for rapid and continuous deployment and allow estimation at smaller geographic scales compared to traditional approaches to data collection, including representative household surveys and retrospective reporting data. These advantages of county-level data in CTIS, compared to the state-level data available from the Census Household Pulse Survey are evident in their respective performance in capturing sub-state heterogeneity in vaccination intentions and coverage. The correlation between parental hesitancy and coverage among children ages 12 to 17 years for CTIS was -0.78, compared to a correlation coefficient of -0.44 for ASPE estimates based on the Census Household Pulse Survey.^5^

Beyond the COVID-19 pandemic, large-scale, low-cost surveys could be applied to regularly generate and update estimates of county-level geographic heterogeneity in determinants of health, healthcare access, and health outcomes. Designing integrated health measurement systems that intentionally combine sources with different advantages across the spectrum of timeliness, geographic granularity, and representativeness can maximize the benefits of data collection relative to their costs. Future large-scale, low-cost data collection efforts should ensure sufficient indicators are incorporated in the survey instrument for post-stratification, as well as availability of appropriate reference indicators for secondary bias-adjustment.

The results of our study should be interpreted in the context of several limitations. First, to predict plateau coverage for children ages 5 to 11 years we assumed that the relationship between hesitancy and coverage observed for children ages 12 to 17 applies to this younger age group. Our three-month validation supports this assumption, which is necessary for prospective estimation. Second, estimates of hesitancy for children of different age groups only became available in Wave 12 of the CTIS survey, and respondents are only asked about intentions to vaccinate their oldest child. Third, we rely on historical relationships between hesitancy and observed coverage, which will not capture the evolving COVID-19 policy and epidemiologic landscape. Fourth, our analytic framework is designed to capture geographic variation in coverage but not variation by other important population characteristics such as race/ethnicity within small geographic areas. Despite these limitations, our estimates reflect a principled approach to generating bias-adjusted estimates of vaccination coverage that can be used to inform decisions and evaluate actual progress against a reference scenario.

## Conclusion

A combination of post-stratification and secondary normalization to an unbiased reference can reduce bias in large-scale, low-cost survey data. Applying this method to predict long-term county-level COVID-19 vaccination coverage among children ages 5 to 11 years, we find substantial sub-state geographic heterogeneity and disparities in the pace of scale-up. Although direct estimates of vaccination coverage from the COVID-19 Trends and Impact Survey are biased, a multi-step regression strategy can result in bias-adjusted actionable predictions on the time-scale and geographic-scale required for proactive decision-making in the pandemic.

## Supporting information

Supplemental Results

## Data Availability

Survey microdata are not publicly available because survey participants only consented to public disclosure of aggregate data, and because the legal agreement with Facebook governing operation of the survey prohibits disclosure of microdata without confidentiality protections for respondents. Deidentified microdata are available to researchers under a Data Use Agreement that protects the confidentiality of respondents. Access can be requested online (https://cmu-delphi.github.io/delphi-epidata/symptom-survey/data-access.html). Requests are reviewed by the Carnegie Mellon University Office of Sponsored Programs and Facebook Data for Good.

## Acknowledgements

We would like to thank the Delphi Group at Carnegie Mellon University for their role in managing the COVID-19 Trends and Impact Survey. We appreciate feedback from members of the SC-COSMO group (https://sc-cosmo.org/) and Prevention Policy Modeling Lab (https://ppml.stanford.edu/) on this work.

