## Supplemental Results for "Bias-adjusted predictions of county-level vaccination coverage from the COVID-19 Trends and Impact Survey"

**Supplemental Figure S1:** Flowchart for creating the COVID-19 Trends and Impact Survey analytic dataset.

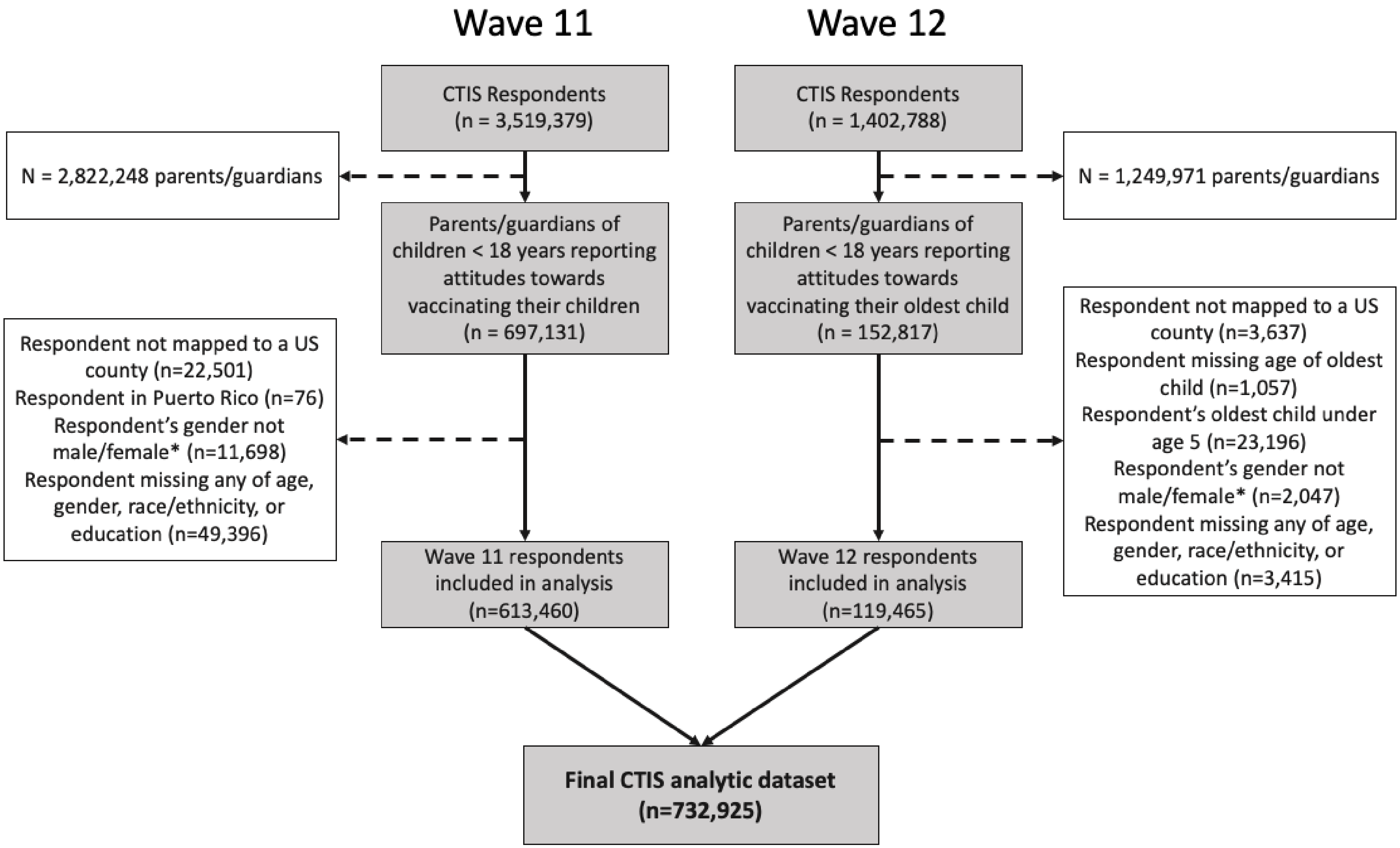


*Analysis could only incorporate respondents reporting their gender as either male or female to match categories available in the American Community Survey for post-stratification.

**Supplemental Figure S2:** Sample size of the analytic sample (n=732,925) from the COVID-19 Trends and Impact Survey, by county.

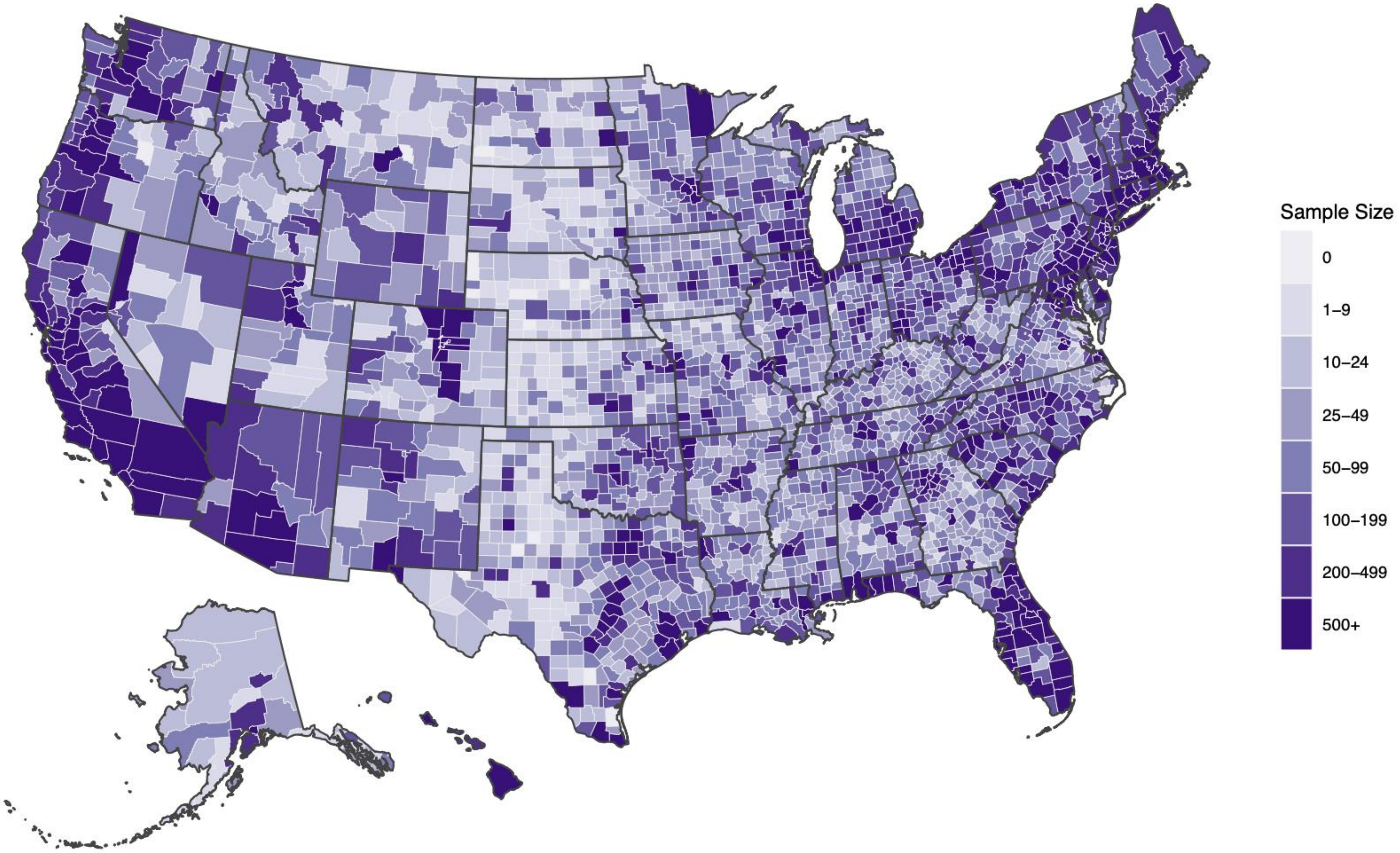

**Supplemental Figure S3:** Sample rate of the analytic sample (n=732,925) from the COVID-19 Trends and Impact Survey, per 1,000 population, by county. Population calculated based on number of children ages 5 to 17 years.

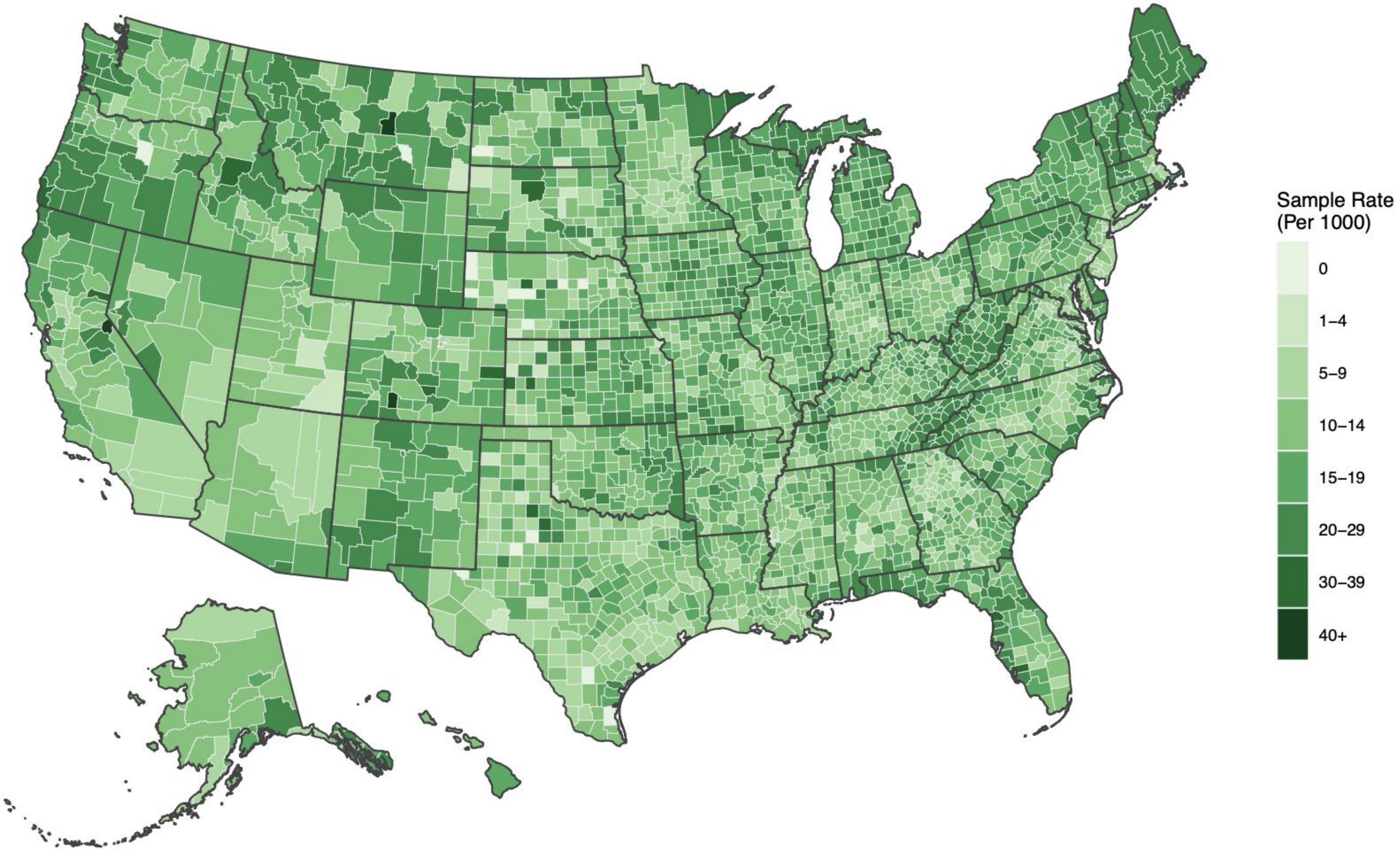

**Supplemental Figure S4:** Comparison of the correlation of county-level estimated hesitancy from CTIS and ASPE and observed complete vaccination coverage on February 1, 2022, for children ages 12-17 years.

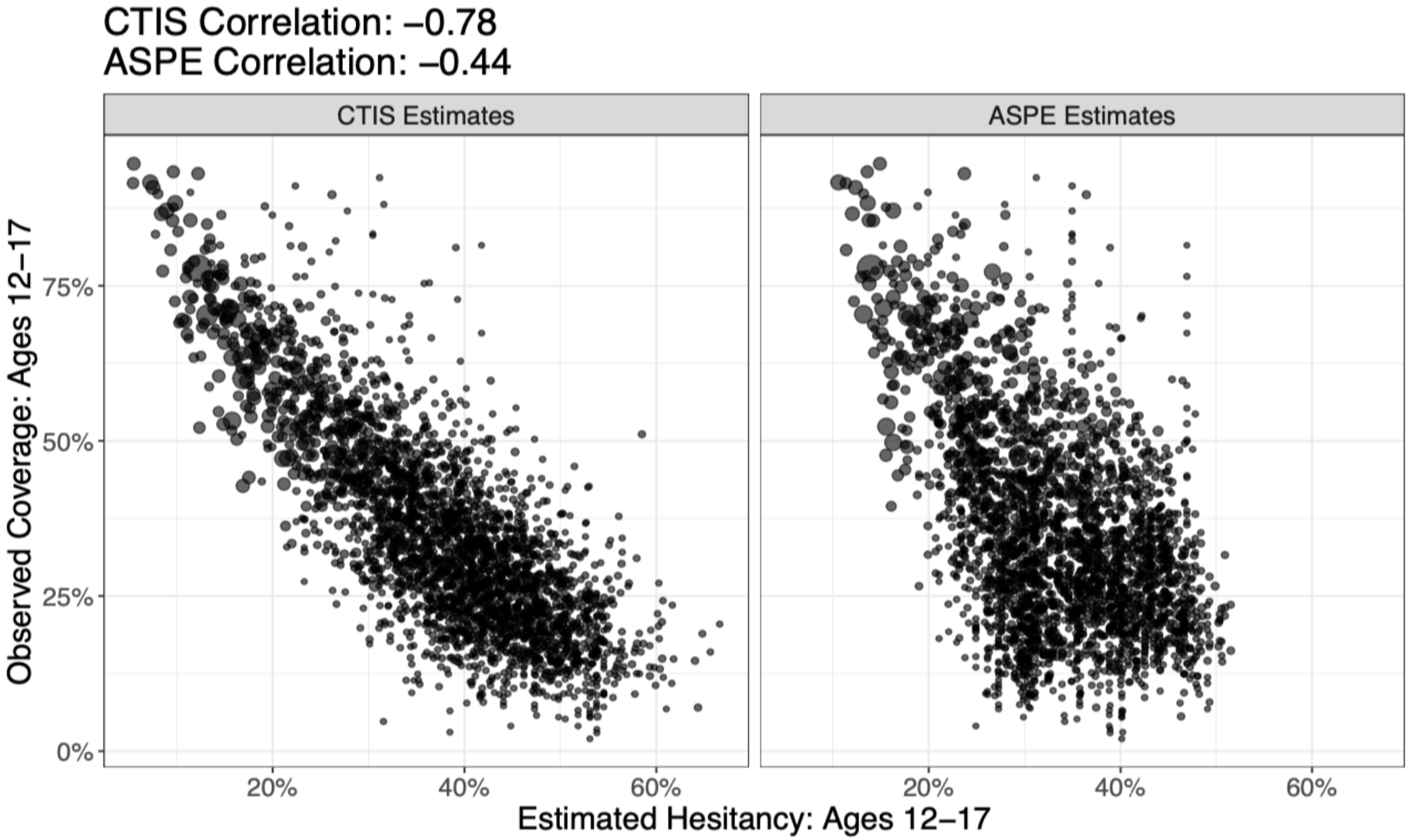

**Supplemental Table S5.** Intraclass correlation coefficient between predicted three-month coverage for children ages 5 to 11 years and observed complete vaccination coverage on February 3, 2022 (three months after Emergency Use Authorization).

| State | Intraclass Correlation |
|---|---|
| **Alabama** | 0.74 |
| **Alaska** | 0.31 |
| **Arizona** | 0.65 |
| **Arkansas** | 0.72 |
| **California** | 0.93 |
| **Colorado** | 0.51 |
| **Connecticut** | 0.86 |
| **Delaware** | 0.96 |
| **Florida** | 0.84 |
| **Idaho** | 0.44 |
| **Illinois** | 0.79 |
| **Indiana** | 0.80 |
| **Iowa** | 0.74 |
| **Kansas** | 0.75 |
| **Kentucky** | 0.72 |
| **Louisiana** | 0.78 |
| **Maine** | 0.92 |
| **Maryland** | 0.82 |
| **Massachusetts** | 0.74 |
| **Minnesota** | 0.62 |
| **Mississippi** | 0.44 |
| **Missouri** | 0.84 |
| **Montana** | 0.40 |
| **Nebraska** | 0.77 |

| State | Intraclass Correlation |
|---|---|
| **Nevada** | 0.72 |
| **New Hampshire** | 0.24 |
| **New Jersey** | 0.79 |
| **New Mexico** | 0.78 |
| **New York** | 0.41 |
| **North Carolina** | 0.71 |
| **North Dakota** | 0.40 |
| **Ohio** | 0.70 |
| **Oklahoma** | 0.71 |
| **Oregon** | 0.76 |
| **Pennsylvania** | 0.83 |
| **Rhode Island** | 0.31 |
| **South Carolina** | 0.59 |
| **South Dakota** | 0.44 |
| **Tennessee** | 0.71 |
| **Texas** | 0.65 |
| **Utah** | 0.63 |
| **Washington** | 0.71 |
| **West Virginia** | 0.63 |
| **Wisconsin** | 0.71 |
| **Wyoming** | 0.49 |
